# Genotyping TOMM40’523 Poly-T Polymorphisms Using Whole-Genome Sequencing

**DOI:** 10.1101/2025.04.23.25326276

**Authors:** Ricardo A Vialle, Lei Yu, Yan Li, Roberto T Raittz, Jose M Farfel, Philip L De Jager, Julie A Schneider, Lisa L Barnes, Shinya Tasaki, David A Bennett

**Author notes:** **Corresponding author** Ricardo A Vialle, Instructor, Rush Alzheimer’s Disease Center, Rush University Medical Center, Jelke Building, 1750 W. Harrison St., Suite 1009N, Chicago, IL 60612.

## Abstract

The TOMM40’523 poly-T repeat polymorphism (rs10524523), located in the *TOMM40* gene and in linkage disequilibrium with *APOE*, has been associated with cognitive decline and Alzheimer’s disease (AD) progression. Accurate genotyping of this polymorphism is crucial for understanding its role in neurodegeneration. Challenges in processing whole-genome sequencing (WGS) data traditionally require additional PCR and targeted sequencing assays to genotype these polymorphisms. Here, we introduce a novel computational pipeline that integrates multiple short tandem repeat (STR) detection tools in an ensemble machine learning model using *XGBoost*. This approach leverages STR tool predictions, k-mer counts, and related features to enhance poly-T repeat length estimation. Using a sample of 1,202 participants from four cohort studies, we benchmarked our method against PCR-based measures. Our ensemble model outperformed individual STR tools, improving repeat length estimation accuracy (R^2^ = 0.92) and achieving an accuracy rate of 93.2% with PCR-derived genotypes as the gold standard. Additionally, we validated our WGS-derived genotypes by replicating previously reported associations between TOMM40’523 variants and cognitive decline, demonstrating consistency with prior findings. Our results suggest that computational genotyping from WGS data is a scalable and reliable alternative to PCR-based assays, enabling broader investigations of *TOMM40* variation in studies where WGS data is available.

## Introduction

Late-onset Alzheimer’s disease (LOAD) is a complex neurodegenerative disorder influenced by multiple factors. The *apolipoprotein E* (*APOE*) ε4 allele is the most well-established genetic risk factor for LOAD. Located in the same locus and in linkage disequilibrium (LD) with *APOE*, there is the *translocase of outer mitochondrial membrane 40* (*TOMM40*) gene. Growing evidence highlights that a variable-length poly-T repeat polymorphism (rs10524523) located in the intron 6 of *TOMM40*, referred to as TOMM40’523, is a key contributor to age at onset and AD progression (Roses 2010; Johnson et al. 2011; Chiba-Falek, Gottschalk, and Lutz 2018). This polymorphism is usually categorized into three repeat length groups: short (‘523-S, ≤19 repeats), long (‘523-L, 20–29 repeats), and very long (‘523-VL, ≥30 repeats) (Roses 2010). These classifications derive from studies linking *TOMM40* repeat lengths to neuropathological changes and cognitive outcomes in AD (Johnson et al. 2011; Chiba-Falek, Gottschalk, and Lutz 2018). We previously reported that, among whites, almost all ε4 carriers also present the ‘523-L allele, and almost all the non-ε4 carriers were absent of the ‘523-L allele, while in African Americans, the LD is much weaker and less than half of the ε4 carriers carry the ‘523-L (Yu, Lutz, Wilson, Burns, Roses, Saunders, Yang, et al. 2017). This heterogeneity suggests that *TOMM40* variation may partly account for ancestry differences in AD susceptibility and the inconsistent effect size of *APOE* ε4 across diverse populations (Bussies et al. 2020). Among white *APOE* ε3 homozygotes, ‘523-S/S genotypes display a faster decline in global cognition compared to individuals with ‘523-S/VL or ‘523-VL/VL (Yu, Lutz, Wilson, Burns, Roses, Saunders, Gaiteri, et al. 2017). Further, the association of ‘523-L with cognitive decline is attenuated after controlling for neuropathologies (Yu, Lutz, Farfel, Wilson, Burns, Saunders, De Jager, et al. 2017), consistent with partial mediation.

We are unaware of any commercial laboratories that perform high-throughput TOMM40 genotyping. Thus, people have turned to leveraging the widespread availability of whole-genome sequencing (WGS). Genotyping the *TOMM40* poly-T repeat from WGS data is technically challenging due to its repetitive sequence and high homology, usually requiring additional targeted Sanger sequencing or PCR-based assays (Roses 2010; Linnertz et al. 2012). Many efforts have focused on developing computational tools to accurately genotype short tandem repeats (STR) from WGS data (Oketch, Wain, and Hollox 2024). However, it’s unknown how they perform in the case of *TOMM40* poly-T repeat relative to the gold-standard PCR. Here, we benchmarked these tools against PCR-based measurements and found that they may produce inaccurate poly-T length estimations or fail to provide results in some cases. Therefore, we propose a new machine-learning ensemble tool specifically designed to improve the *TOMM40* poly-T genotyping using short-read WGS. We validate our tool by replicating previous results linking specific *TOMM40* poly-T genotypes to cognitive decline and neuropathological indices of AD (Yu, Lutz, Farfel, Wilson, Burns, Saunders, De Jager, et al. 2017; Yu, Lutz, Wilson, Burns, Roses, Saunders, Gaiteri, et al. 2017).

## Methods

### Study participants

Participants came from multiple ongoing longitudinal cohort studies conducted by the Rush Alzheimer’s Disease Center (RADC). Participants came from the Religious Orders Study (ROS) and the Rush Memory and Aging Project (MAP), while a smaller number were from the Minority Aging Research Study (MARS) and the African American Clinical Core (AA Core) (Marquez et al. 2020; David A. Bennett et al. 2018). All studies are prospective, community-based investigations of risk factors for cognitive decline, incident Alzheimer’s disease (AD) dementia, and other health outcomes. ROS has been recruiting Catholic nuns, priests, and brothers from across the United States since 1994, while MAP recruits participants from retirement communities and subsidized senior housing facilities and individual homes throughout Chicago and northeastern Illinois since 1997. All ROS/MAP participants agree to brain donation after death. MARS, launched in 2004, focuses on cognitive decline and AD risk in older Black Americans, with optional brain donation after death. The AA Core, active since 2008, supports research on cognition, dementia risk factors, pathology, neuroimaging, and biomarkers among African Americans, with optional brain donation. All participants are enrolled without known dementia at baseline. Participants undergo annual cognitive and clinical assessments. All studies were approved by an Institutional Review Board of Rush University Medical Center, and all provided written informed consent. An Anatomical Gift Act was provided by each participant who agreed to brain donation. As of early March 2025, 4,052 ROS/MAP participants were enrolled, of whom 2,127 came to autopsy (52.5%); 906 MARS and 435 AA Core participants were enrolled, of whom 134 had an autopsy (10%). The current study focused on deceased individuals with whole-genome sequencing (WGS) data and PCR-derived *TOMM40* poly-T lengths (N=1,202). The mean age at death was nearly 90 years, 2/3rds were female, and the mean education level was 16 years (**Table 1**).

**Table 1.**
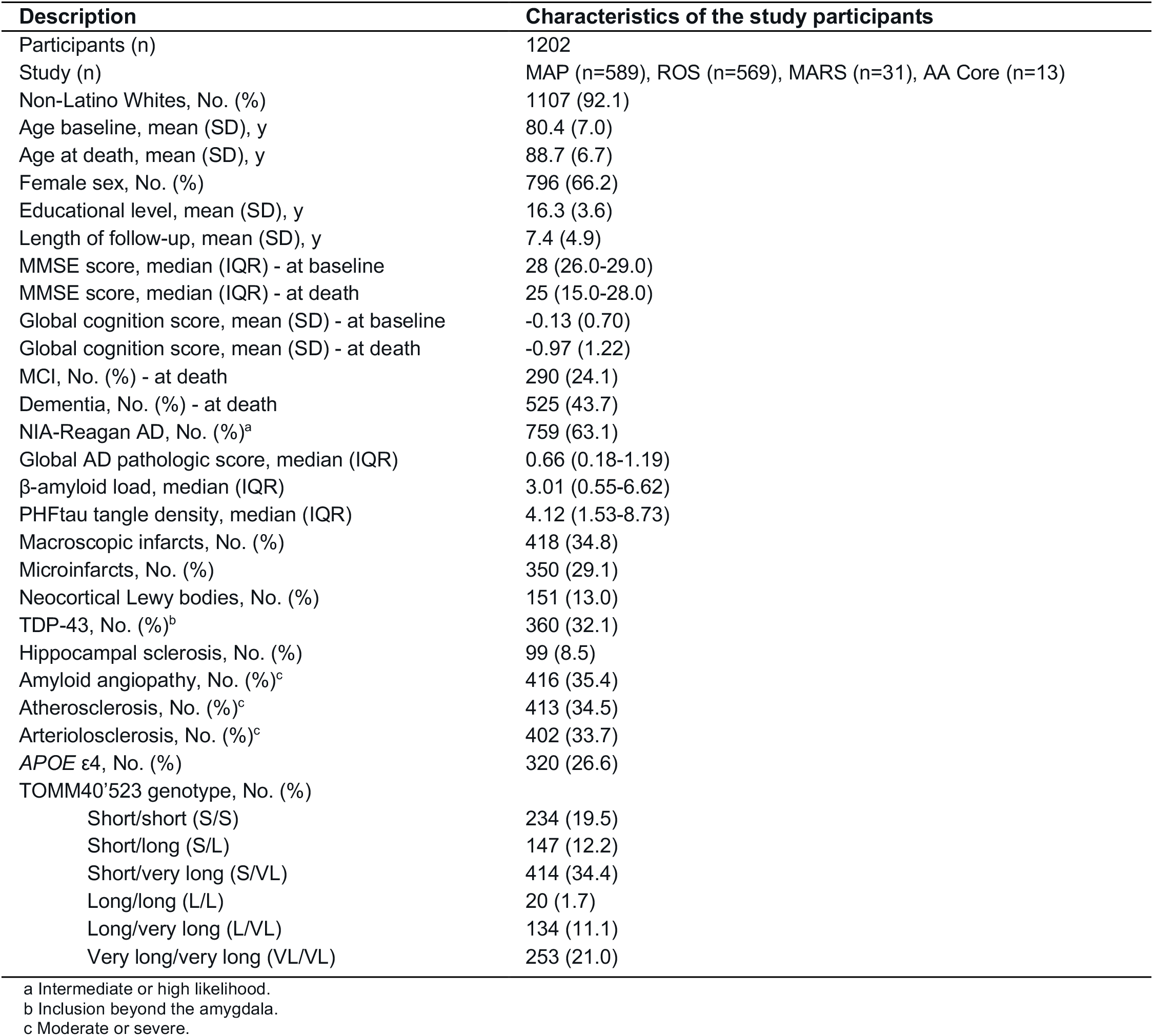
Basic characteristics of the study participants.

### Annual cognitive assessments and postmortem neuropathologic evaluations

Participants underwent uniform annual cognitive assessments for up to 23 years (Mean = 7.4, SD = 4.9). Cognitive performance was assessed using 21 tests, of which 18 generated a global cognitive score and five domains (Boyle et al. 2013). Scores from each test were standardized using the baseline mean and SD. The resulting z-scores were averaged across the tests to obtain composite measures of global cognition and, separately, 5 cognitive domains. These composite measures minimize floor and ceiling effects commonly observed in individual tests.

At autopsy, common age-related neuropathologies were quantified, including Alzheimer’s disease (AD), macroscopic infarcts, microinfarcts, Lewy bodies, hippocampal sclerosis, TDP-43, cerebral amyloid angiopathy (CAA), atherosclerosis, and arteriolosclerosis. Each is briefly described below.

- β-amyloid and phosphorylated PHFtau tangles were assessed in eight brain regions using immunohistochemistry (D. A. Bennett et al. 2005). The percent area positive for β-amyloid was computed for each region using image analysis and averaged across the regions to obtain a summary measure of β-amyloid load. The density of PHFtau tangles per mm^2^ was computed for each region using a stereological mapping station and averaged to obtain a summary measure of PHFtau tangle density.
- Chronic macroscopic infarcts were recorded during gross examination and verified histologically (Schneider et al. 2003). Chronic microinfarcts were identified in at least nine brain regions using hematoxylin and eosin (H&E) staining (Arvanitakis et al. 2011).
- The presence of Lewy bodies in neocortical regions was identified using α-synuclein immunostaining (Schneider et al. 2012).
- Hippocampal sclerosis, defined as severe neuronal loss and astrogliosis of CA1 and/or the subiculum, was determined using H&E staining (Nag et al. 2015).
- TDP-43 pathology was assessed in five brain regions using monoclonal antibodies to phosphorylated TDP-43 and was rated on a four-level scale: no inclusions, inclusions in the amygdala, inclusions in the amygdala and limbic system, or inclusions in the amygdala, limbic system, and neocortex (Yu, De Jager, et al. 2015).
- Cerebral amyloid angiopathy was assessed in four neocortical regions using monoclonal antibodies to β-amyloid (Yu, Boyle, et al. 2015). The amount of β-amyloid deposition in the vessel walls was scored for each region, and the average scores across the regions were summarized into a four-level scale representing none, mild, moderate, or severe.
- Atherosclerosis was assessed in the circle of Willis during gross examination, while arteriolosclerosis was assessed in the anterior basal ganglia using H&E staining. Both were rated on a four-level scale of none, mild, moderate, or severe (Arvanitakis et al. 2016).

### TOMM40’523 PCR genotyping

DNA was extracted from peripheral blood or frozen postmortem brain tissue. Genotyping was performed at Polymorphic DNA Technologies (Alameda, CA), with the vendor blinded to all clinical and neuropathologic information. The ‘523 genotype was determined based on the length of the poly-T repeat, as previously described (Caselli et al. 2012). Briefly, polymorphic simultaneous polymerase chain reaction (PCR) was performed to amplify each polymorphism. Then, a bidirectional direct Sanger sequencing of the DNA templates was performed on an Applied Biosystems 3730xl DNA Analyzer (Applied Biosystems Inc, Carlsbad, CA), followed by sequence data analysis. The measured poly-T lengths were converted to ‘523 short allele (‘523-S) when the repeat length was less than 20 bp, to long allele (‘523-L) when the repeat length was between 20 bp and 29 bp, and to very long allele (‘523-VL) when poly-T repeat length was equal to 30 bp or more.

### Whole-genome sequencing data

Whole-genome sequencing (WGS) data were previously generated from DNA samples from blood or cortex tissues as part of the AMP-AD consortium (Reddy et al. 2024; De Jager et al. 2018). Briefly, for the first WGS release (syn10901595) (De Jager et al. 2018), DNA samples were obtained from the Broad Institute (ProjectDEJ11898). Libraries were prepared using the KAPA Hyper Library Preparation Kit following the manufacturer’s instructions, and sequencing was performed on an Illumina HiSeq X sequencer (v2.5 chemistry) using 2 x 150bp cycles. Raw sequencing reads were realigned to the GRCh38 human reference genome using BWA-MEM. For the second WGS data release (syn51732520) (Reddy et al. 2024), libraries were generated using the Illumina Tru-Seq PCR-Free protocol, followed by short-read sequencing technology with a mean read length of 150 base pairs performed at the New York Genome Center (NYGC). Sequencing reads were aligned to the GRCh38 human reference using the Burrows-Wheeler Aligner (BWA-MEM v0.7.15). CRAM files were obtained for downstream analysis.

### Genotyping TOMM40’523 from WGS

We used a dataset comprising 1,202 whole-genome sequencing (WGS) samples from four cohort studies: ROS, MAP, MARS, and AA Core. All samples had been previously genotyped using PCR, as described in earlier studies (Caselli et al. 2012; Yu, Lutz, Wilson, Burns, Roses, Saunders, Gaiteri, et al. 2017; Yu, Lutz, Farfel, Wilson, Burns, Saunders, De Jager, et al. 2017; Yu, Lutz, Wilson, Burns, Roses, Saunders, Yang, et al. 2017), providing a reference dataset for validating computational predictions. We *in silico* measured poly-T lengths using three well-known STR tools *ExpansionHunter* (Dolzhenko et al. 2019), *HipSTR* (Willems et al. 2017), and *GangSTR* (Mousavi et al. 2019). Customized configurations were necessary to genotype *TOMM40* using STR tools with WGS data. For *ExpansionHunter*, the following configuration was set: VariantType = “Repeat”, LocusStructure = “(T)+”, and ReferenceRegion = “19:44899792-44899826”. For *GangSTR*, the targeted locus was specified as a BED file. For *HipSTR*, in addition to restricting the region to the locus, the parameter *--max-flank-indel* was increased to 0.9, and *--min-reads* was set to 15.

To predict allele-specific TOMM40’523 poly-T lengths, we developed an ensemble machine-learning model using *XGBoost* (Chen and Guestrin 2016). The model incorporates STR tool predictions and additional features extracted from sequencing data. Features included tool-specific VCF file annotations, k-mer counts (3– 50 bp), and neural network-based haplotype predictions. The dataset of 1,202 individuals was split 50/50 for training (n = 600) and testing (n = 602), ensuring a balanced distribution of the six genotypes between partitions using the *groupdata2::partition* function in R. Next, alleles from each set were separated (i.e., 600 × 2 = 1,200 alleles for training and 602 × 2 = 1,204 alleles for testing) and used as features for model training and evaluation.

A set of features was derived from three primary sources. The first source was a table of k-mer counts, with k ranging from 3 to 50. The second source included *ExpansionHunter*-predicted poly-T lengths, along with additional extracted features. The third source contained *GangSTR*-predicted lengths and corresponding supplementary features. Results derived from *HipSTR* were not included because the tool failed to produce results in a few cases. Briefly, k-mer counts were obtained using *Jellyfish* (Marçais and Kingsford 2011), using *STRling* (Dashnow et al. 2022) to extract reads from the *TOMM40* repeat region in the reference genome (chr19:44,899,792–44,899,826; GRCh38). Features from *ExpansionHunter* were extracted directly from the resulting VCF file, yielding nine features, including poly-T lengths for each allele, confidence intervals, number of flanking reads, number of in-repeat reads, number of spanning reads, and others (**Supplementary Table S1**). From *GangSTR*, 18 features were extracted, including allele length measures, read depth, quality score, maximum likelihood, and counts of enclosing, spanning, fully repetitive, and bounding reads (**Supplementary Table S1**).

We then generated additional features by training multi-layer perceptron (MLP) neural networks using only k-mer (3-mer to 50-mer) counts as input. First, a neural network was trained to predict the six-category genotypes (‘523-S/S, ‘523-S/L, ‘523-S/VL, ‘523-L/L, ‘523-L/VL, and ‘523-VL/VL). Model training was conducted in R using the *parsnip::mlp* function in classification mode with the *nnet* engine, allowing a maximum of 100,000 weights. The parameters were set as follows: L2 regularization penalty = 0.666, 100 million epochs, and 10 hidden neurons. The same 50/50 training and testing split datasets used for the complete model were applied. The classifier achieved an accuracy of 0.96 (kappa = 0.95) on the training set and 0.91 (kappa = 0.89) on the test set. In addition, three MLP models were trained to classify haplotypes into three independent categories: S (i.e., ‘523-S/S, ‘523-S/L, ‘523-S/VL vs. others), L (i.e., ‘523-L/L, ‘523-S/L, ‘523-L/VL vs. others), and VL (i.e., ‘523-VL/VL, ‘523-S/VL, ‘523-L/VL vs. others). The same network parameters and inputs were applied. Individual classifiers achieved 0.99, 0.95, and 0.96 accuracies for predicting ‘523-S, ‘523-L, and ‘523-VL groups in the test set, respectively. The final model included 10 additional features: the predicted genotype class for six categories, the corresponding probabilities for each of those six classes, and the predicted probabilities from three haplotype classifiers. The final model included 10 additional features: the predicted genotype class (one feature), the corresponding probabilities for each of those six classes (6 features), and the predicted probabilities from three haplotype classifiers (3 features).

The XGBoost regression model was trained using 85 features. Hyperparameter tuning was performed using the *mlr* R package, with a 3-fold cross-validation and 96 random search iterations. The trained model was evaluated using the root mean squared error (RMSE). Final optimized parameters were set as follows: objective = reg:squarederror, eval_metric = rmse, nrounds = 300, eta = 0.0382, booster = gbtree, nthread = 1, alpha = 0, lambda = 2.83, max_depth = 5, min_child_weight = 11.5, subsample = 0.68, colsample_bytree = 0.488, gamma = 15.2, grow_policy = lossguide. Performance was of 0.99 correlation and 1.32 RSME in the training set, and 0.96 correlation and 2.45 RSME in the test set.

### Analyses

Multivariable linear or logistic regression analyses were applied to test for the association of TOMM40’523 genotypes with cognitive or neuropathologic measures depending on the distribution of the outcome variables (**Supplementary Table S2**). In each of these models, the individual cognitive or neuropathologic measure was the outcome, and TOMM40’523 genotypes (e.g., ‘523-S/S vs. others, or ‘523-S/S vs. ‘523-S/VL and ‘523-VL/VL) were the predictors while controlling for age at death and sex. For the associations with cognitive decline and cognitive resilience, person-specific slopes of change derived from linear mixed-effects models were used as the outcomes (De Jager et al. 2012; Oveisgharan et al. 2023). Considering a total of 13 cognitive and neuropathologic indices were tested, Bonferroni correction was used to control for the inflation of Type-1 error. The analyses were performed using R 4.1.2, with functions *stats::lm, stats::glm,* and *ordinal::clm*.

### Implementation

The prediction tool was implemented using the Snakemake (Mölder et al. 2021) and Bioconda (Grüning et al. 2018) frameworks to manage the installation and execution of the tools. The mandatory requirements are a Unix environment with Python, Conda/Mamba, and Git. The Snakemake pipeline manages tools execution in modules, starting by extracting reads from a WGS alignment file (BAM or CRAM format), execution of standalone STR tools and k-mer extraction/counting to generate required features, and ending with TOMM40’523 poly-T lengths prediction and genotype reconstruction (**Supplementary Figure S1**).

## Results

### Benchmarking poly-T length with STR methods

We first evaluated the performance of short tandem repeats (STR) genotyping tools to estimate TOMM40’523 poly-T lengths and respective haplotypes from short-read whole-genome sequencing (WGS). We tested three well-known STR tools, *ExpansionHunter, HipSTR*, and *GangSTR,* on 1,202 WGS data and assessed their performance against the PCR-genotyped reference dataset. In relation to defining the six genotyping categories, *ExpansionHunter* achieved the highest accuracy rate of 93.09% with a Cohen’s kappa value of 0.91, followed by *HipSTR* with 88.52% accuracy and a kappa value of 0.88, and *GangSTR* with an accuracy rate of 63.06% and a kappa value of 0.54 (**Supplementary Figure S2A-I**). Despite good overall performance, the STR tools struggled to accurately identify particular genotypes such as ‘523-L/L, with *ExpansionHunter* and *HipSTR* achieving accuracies below 40%. We further examined the accuracy of each tool in estimating the actual poly-T repeat lengths by comparing WGS-derived and PCR-measured lengths (**Figure 1A**). *ExpansionHunter* tended to overestimate repeat lengths, resulting in a moderate correlation (R^2^ = 0.55). *GangSTR* showed an even weaker correlation (R^2^ = 0.44), with a tendency to underestimate the repeat lengths. By contrast, *HipSTR* exhibited the most reliable tool for estimating repeat length with the strongest correlation with PCR-derived measures (R^2^ = 0.89). However, *HipSTR* failed to produce results in 4.3% of samples tested while the other two tools produced results in all samples.

**Fig. 1.**
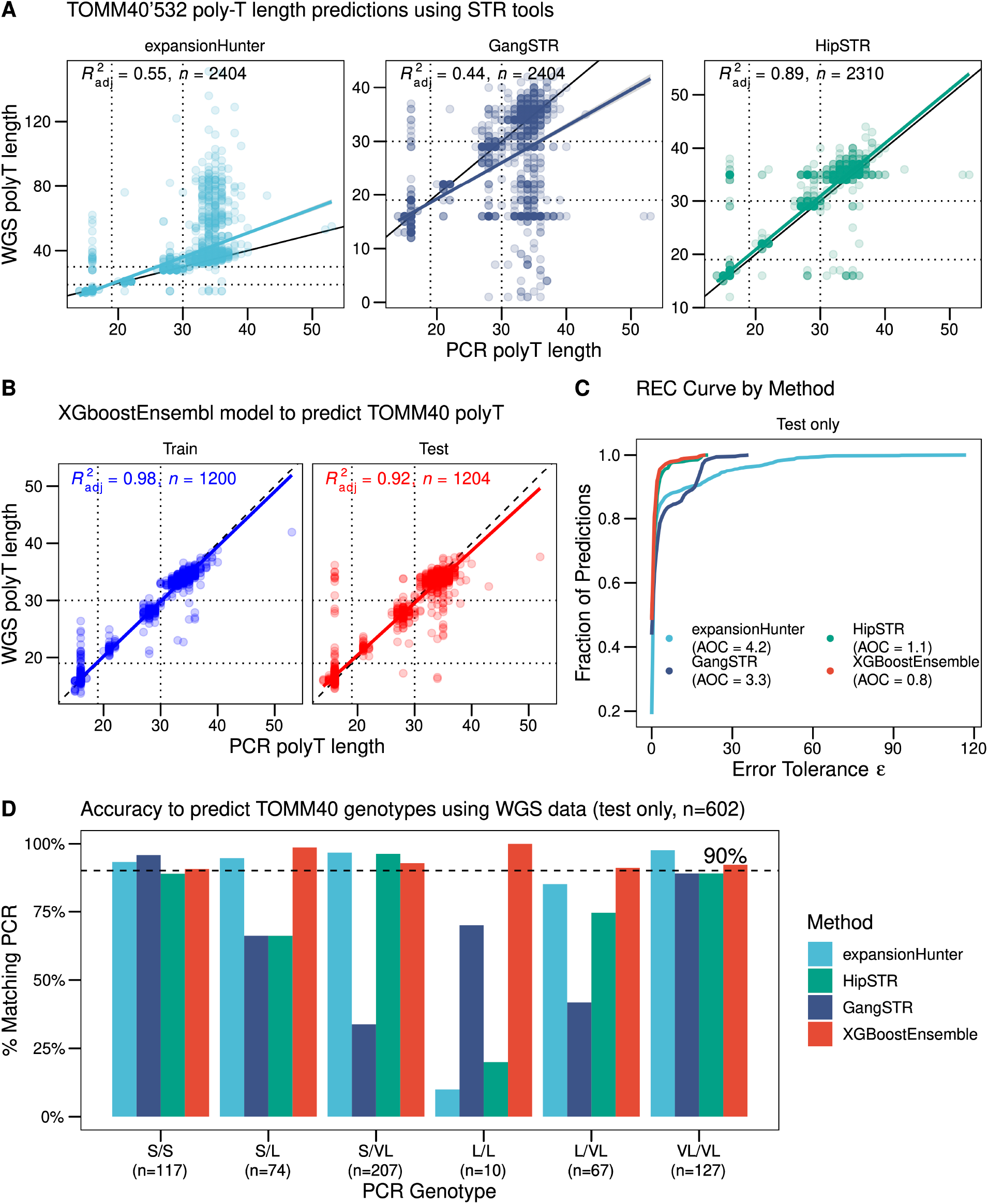
Benchmarking TOMM40’523 measures using WGS data. **A)** Comparison between PCR-derived poly-T lengths (x-axis) and measures given by STR tools using WGS data (y-axis). **B)** Results for *XGBoost-predicted* poly-T lengths (y-axis) and PCR poly-T lengths (x-axis). Results are split by Train and Test samples. Dotted lines, in **A** and **B**, indicate repeat lengths equal to 19 and 30 for PCR-and WGS-derived measures representing the separation of haplotypes (‘523-S, ≤19 repeats, ‘523-L, 20–29 repeats, ‘523-VL, ≥30 repeats) **C)** Regression error characteristics (REC) curve of poly-T length performances comparison between STR tools and *XGBoost* predictions, showing Test samples only (n=602). **D)** Accuracy measured as % of genotyping group classification compared to PCR-derived genotypes (‘523-S/S, ‘523-S/L, ‘523-S/VL, ‘523-L/L, ‘523-L/VL, and ‘523-VL/VL). The dashed line indicates 90% matching. Each color represents one method. Results are shown for Test samples only (n=602).

### *XGBoost* ensemble model improves poly-T length accuracy

To improve TOMM40’523 poly-T length estimations derived from WGS data, we developed a machine learning prediction model based on a gradient-boosting regression model (via *XGBoost*) incorporating 85 features derived from *ExpansionHunter* and *GangSTR,* along with a set of features based on k-mer fragments extracted from the locus (see Methods for details). The dataset was split 50/50 for training and testing. As a result, the prediction model demonstrated an improvement in TOMM40’523 poly-T length estimation, achieving an R^2^ of 0.92 on the test set (R^2^ = 0.97 on the training set) (**Figure 1B**), surpassing the best-performing individual tool, *HipSTR* (R^2^ = 0.89). Improvements were also reflected in better regression error characteristics (REC) curves (**Figure 1C**). When these refined length estimates were converted back into genotype groups, the *XGBoost* ensemble model outperformed individual STR tools, achieving an accuracy rate of 93.19% with a kappa value of 0.91, closely mirroring *ExpansionHunter*’s performance (**Figure 1D; Supplementary Figure S2J-L**). More importantly, the accuracy across all genotype groups consistently exceeded 90%, representing a notable improvement, particularly for the challenging homozygous long alleles (i.e., ‘523-L/L). Further, results were obtained in all subjects.

### Replication analysis of prior findings

To assess whether the discordance between WGS and PCR-based genotyping was due to random variation or systematic bias, we examined potential discrepancies in the context of known linkage disequilibrium (LD) patterns. Importantly, we included only test-set samples in this analysis to avoid bias from the training model. The long haplotype (‘523-L) is expected to be strongly linked to *APOE* ε4 in non-Latino Whites (Yu, Lutz, Wilson, Burns, Roses, Saunders, Yang, et al. 2017). However, the concordance between PCR and WGS-derived genotypes was lower in these cases when using our prediction model (**Supplementary Table S3**). While for PCR-genotyped samples, 97.9% of ‘523-L’ alleles were also ε4 carriers, the proportion observed in WGS-based calls was 87.9%. By contrast, the percentage of ε4 carriers with ‘523-L was comparable across both genotyping methods (∼94%). In addition, no significant discrepancies were observed in non-Latino Black individuals, with about half of ε4 carriers carrying the ‘523-L allele (**Supplementary Table S4**).

We then sought to replicate prior findings from two studies using the WGS-inferred genotypes. The first analysis focused on the association between ‘523-L and measures of cognitive decline, cognitive resilience and neuropathologic indices (Yu, Lutz, Farfel, Wilson, Burns, Saunders, De Jager, et al. 2017). We found consistent results between PCR- and WGS-derived genotypes. As previously reported, results based on WGS showed that long alleles were associated with faster cognitive decline and multiple neuropathologies, including β-amyloid, tangles, TDP-43, and CAA. In addition, there was no association of ‘523-L derived from PCR and WGS with cognitive resilience (i.e., residual cognitive decline after accounting for neuropathologic indices), confirming earlier findings (**Figure 2A**). Among NLBs, due to the limited sample size, only an association of WGS-derived ‘523-L with CAA was found. Importantly, the effect sizes for the WGS-derived ‘523-L were comparable to those for the PCR-derived genotypes (**Supplementary Figure S3**). The second analysis aimed to replicate findings in *APOE* ε3 homozygotes, where prior studies suggested an association between short alleles and cognitive decline compared to ‘523-S/VL and ‘523-VL/VL genotypes (Yu, Lutz, Wilson, Burns, Roses, Saunders, Gaiteri, et al. 2017). For these tests, given the limited sample size, we combined both training and test samples. Once again, the results were very similar between PCR and WGS-based genotypes (**Figure 2B**), suggesting that any residual discrepancies, such as the previously mentioned ‘523-L vs. ε4, are unlikely to impact association studies, particularly in well-powered datasets typical of WGS studies. These findings support the robustness of WGS-derived genotypes for replication studies and reinforce prior associations between *TOMM40* polymorphisms and cognitive outcomes.

**Fig. 2.**
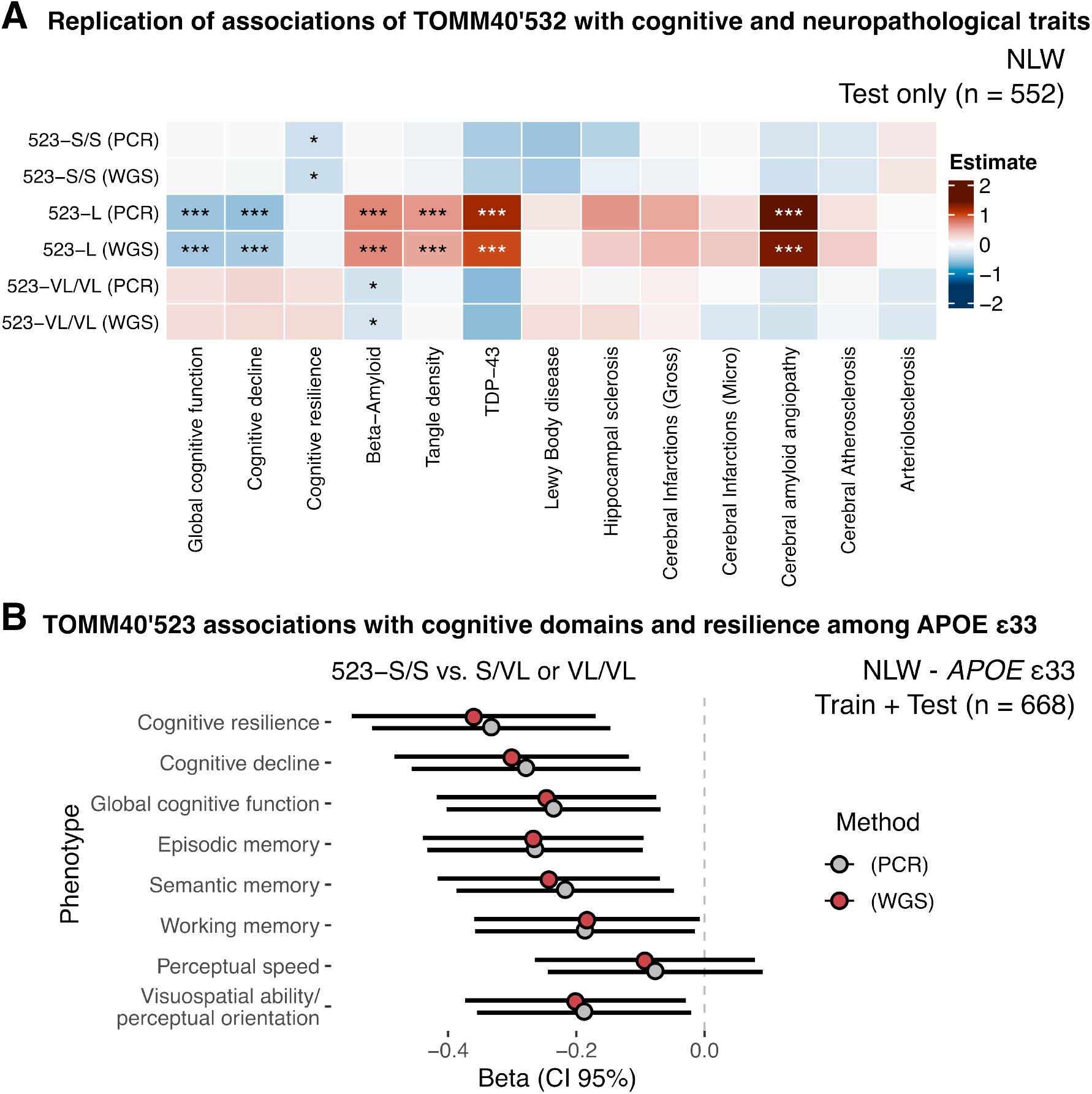
Replication of TOMM40’523 associations with cognitive and neuropathological indices. **A)** Heatmap showing associations of both PCR- and WGS-derived (*XGBoost*) *TOMM40* genotypes versus different cognition and neuropathologic indices. Color indicates the strength of association (estimate). Asterisks indicate Bonferroni adjusted *P*-values: \**P* ≤ 0.05, \*\**P* ≤ 0.01, \*\*\**P* ≤ 0.001. Only included NLWs and Test samples (n=552). **B)** Beta and 95% confidence interval of associations of cognitive phenotypes between participants with ‘523-S/S versus ‘523-S/LV or ‘523-VL/VL. Only included NLWs and *APOE* ε3 homozygotes. Train and Test samples were considered (n=668). All tests are adjusted by age at death and sex.

## Discussion

Our findings underscore the potential of computational approaches for accurately genotyping TOMM40’523 polymorphisms using WGS data. By integrating multiple STR detection tools with an ensemble machine learning model, we improved on the existing methods, particularly in genotyping challenging haplotypes such as ‘523-L/L’. The proposed approach achieved high concordance rates with PCR-based assays and replicated results from PCR polymorphisms, demonstrating its viability as a scalable approach for large-scale genetic studies.

Among individual STR tools, *HipSTR* displayed the best overall performance in measuring poly-T lengths and genotype categories. *HipSTR,* however, failed to deliver results for nearly 5% of samples analyzed. *ExpansionHunter*, although it resulted in the best genotype category classification, tended to overestimate lengths (above VL categories) while *GangSTR* provided underestimated lengths. Our ensemble modeling proved to be particularly valuable for resolving challenging genotype calls, especially for the ‘523-L/L genotypes where individual tools showed the poorest performance. By combining predictions from multiple sources through the *XGBoost* model, we achieved consistent accuracy exceeding 90% across all genotype categories. This represents a substantial improvement over individual tools and demonstrates the value of integrating complementary approaches for complex genetic variants.

The successful replication of previously established associations between *TOMM40* poly-T genotypes and cognitive outcomes validates the utility of our computational method. We confirmed the association of ‘523-L alleles with faster cognitive decline and its attenuation after accounting for neuropathological indices, as well as the relationship between short alleles and cognitive decline in *APOE* ε3 homozygotes. These findings suggest that despite some residual discrepancies between PCR and WGS-based genotyping, our method is sufficiently accurate for population-level genetic studies. The study also has limitations. Despite these promising results, future work should focus on benchmarking this method against long-read sequencing data, which could offer higher-resolution insights into poly-T repeat length variation. Further validation across diverse populations is also critical to ensure its generalizability in genetic studies.

## Data Availability

No sequencing data was generated as part of this current study. All relevant codes used in this study are publicly available on GitHub (https://github.com/RushAlz/TOMM40_WGS). Raw sequencing data and individual phenotype resources can be requested at https://www.radc.rush.edu.

https://github.com/RushAlz/TOMM40_WGS

## Declarations

### Ethics approval and consent to participate

The ROS, MAP, MARS and AA Core studies were approved by an Institutional Review Board (IRB) of Rush University Medical Center, Chicago, IL. All participants included in this study agreed to annual clinical evaluation and signed an informed consent and an Anatomic Gift Act agreeing to post-mortem brain donation. All procedures performed in studies involving human participants were in accordance with the ethical standards of the Institutional Review Board of Rush University Medical Center and with the 1964 Helsinki Declaration and its later amendments or comparable ethical standards. Each participant signed an informed consent form to participate in the study.

### Consent for publication

Not applicable.

### Competing interests

The authors declare that they have no competing interests.

### Funding

This work has been supported by the following National Institute on Aging (NIA) grants: P30AG10161 (DAB), P30AG72975 (JAS), R01AG15819 (DAB), R01AG17917 (DAB), U01AG46152 (PLD, DAB), U01AG61356 (PLD, DAB), and R01AG22018 (LLB). The funders had no role in the study design, data collection and analysis, decision to publish, or preparation of the manuscript.

### Authors’ contributions

RAV performed the main analysis, drafted the manuscript, and designed the project. ST, LY, YL and RTR assisted on the analysis. ST, JMF and DAB edited the manuscript and assisted in project design. LLB, JAS, PLD and DAB funded the project. All authors read and approved the final manuscript.

## Acknowledgments

We express our gratitude to the participants and staff of the ROS, MAP, MARS, and AA Core cohorts for their invaluable contributions. We also thank the investigators and staff at the Rush Alzheimer’s Disease Center for their expertise and support in this project.

## Notes

### Competing Interest Statement

The authors have declared no competing interest.

