## Supplementary Material for "Genotyping TOMM40’523 Poly-T Polymorphisms Using Whole-Genome Sequencing"

[2] Graduate Program in Bioinformatics, Professional and Technical Education Sector (SEPT), Universidade Federal do Paraná (UFPR), Curitiba, Paraná, Brazil

[3] Department of Biochemistry, Universidade Federal do Paraná (UFPR), Curitiba, Paraná, Brazil

[4] Center for Translational and Computational Neuroimmunology, Department of Neurology, Columbia University Irving Medical Center, New York, NY, USA

### Supplementary figures

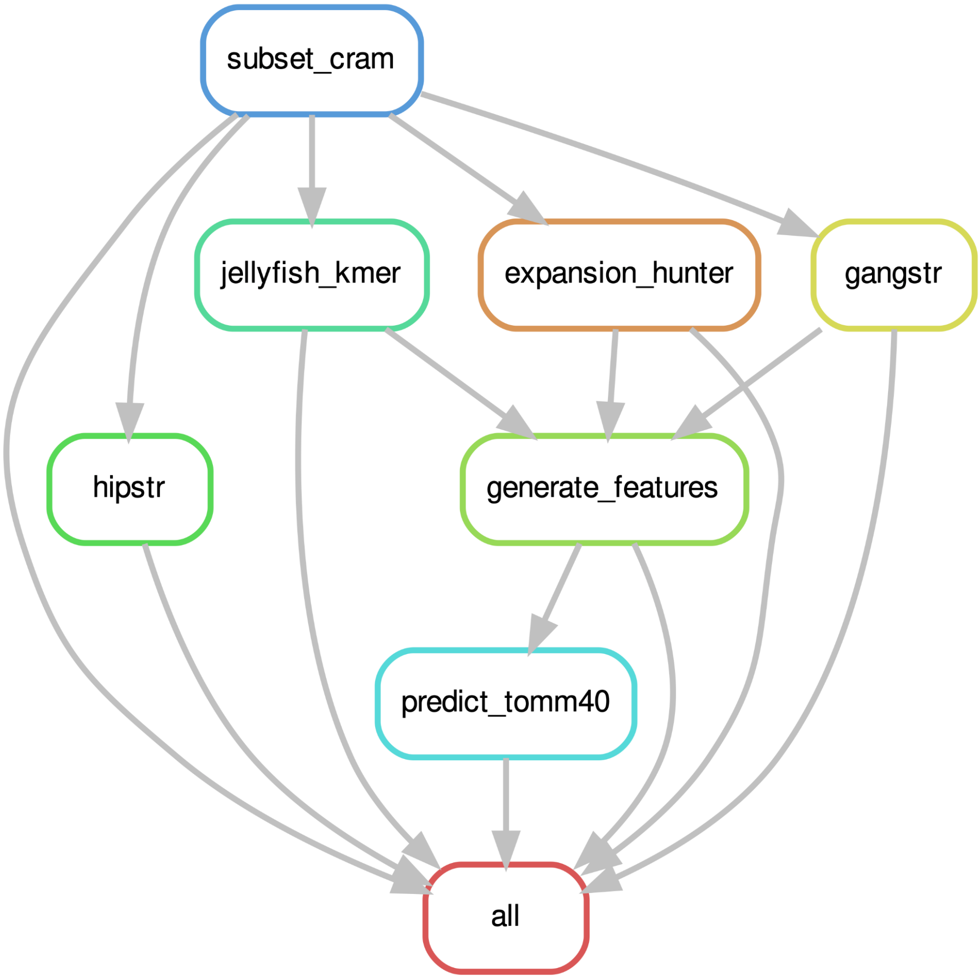


**Supp. Fig. S1. Snakemake pipeline directed acyclic graph (DAG).** The *tomm40_wgs* pipeline receives one or more WGS alignment files (CRAM or BAM) as input. The pipeline first subsets the WGS files to the TOMM40 region, then runs standalone STR genotyping tools (ExpansionHunter, GangSTR, and optionally HipSTR) in parallel, processes k-mer counts, builds a table of derived features, and predicts the TOMM40'523 poly-T lengths. The output file also includes results from individual STR tools and the derived 6-class genotyping.

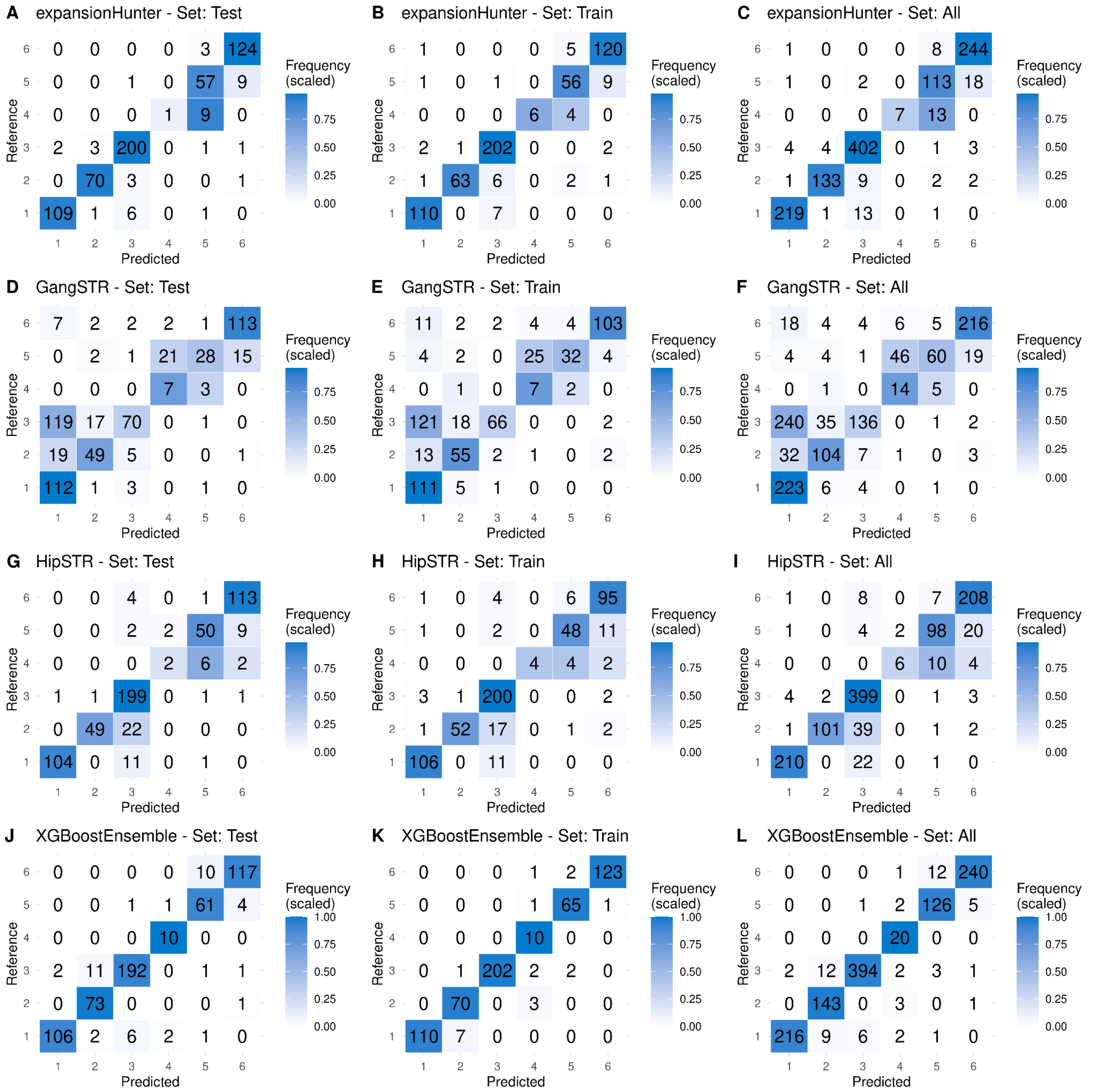


**Supp. Fig. S2. Confusion matrices of TOMM40'523 genotype prediction.** Results for 6-class genotyping (1: 523'-S/S; 2: 523'-S/L; 3: 523'-S/VL; 4: 523'-L/L; 5: 523'-L/VL; 6: 523'-VL/VL) for each STR tool tested and the XGBoostEnsemble prediction model. Results are shown for the testing (Test) and training (Train) subsets used in the creation of the prediction model, as well as for the complete dataset (All). Colors represent the scaled frequency in each genotype class.

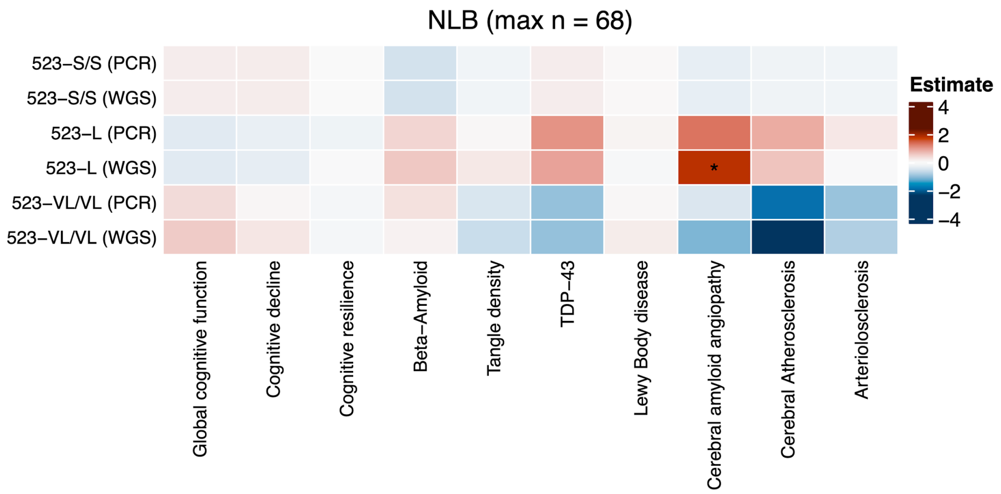


**Supp. Fig. S3. TOMM40'523 associations with cognitive and neuropathological indices among NLBs.** Heatmap showing associations of both PCR- and WGS-derived (*XGBoost*) *TOMM40* genotypes versus different cognition and neuropathologic indices among NLBs (n=68). Color indicates the strength of association (estimate). Asterisks indicate Bonferroni adjusted *P*-values: **P* ≤ 0.05, ***P* ≤ 0.01, ****P* ≤ 0.001. Training and test samples were considered.

### Supplementary tables

**Supp. table S1. Description of features included in the XGBoost regression model**

| feature | source | description |
|---|---|---|
| expansionHunter | expansionHunter | poly-T length predicted by expansionHunter (allele 1) |
| gt_ADFL_expansionHunter | expansionHunter | Number of flanking reads consistent with the allele 1 |
| gt_ADIR_expansionHunter | expansionHunter | Number of in-repeat reads consistent with the allele 1 |
| gt_ADSP_expansionHunter | expansionHunter | Number of spanning reads consistent with the allele 1 |
| gt_REPCI_1_expansionHunter | expansionHunter | Confidence interval for REPCN (lower) (allele 1) |
| gt_REPCI_2_expansionHunter | expansionHunter | Confidence interval for REPCN (upper) (allele 1) |
| gt_REPCN_expansionHunter | expansionHunter | Number of repeat units spanned by the allele 1 |
| gt_SO_expansionHunter | expansionHunter | Type of reads that support the allele; can be SPANNING, FLANKING, or INREPEAT (allele 1) |
| otherAexpansionHunter | expansionHunter | poly-T length predicted by expansionHunter (allele 2) |
| GangSTR | GangSTR | poly-T length predicted by GangSTR (allele 1) |
| gt_REPCN_GangSTR | GangSTR | Genotype given in number of copies of the repeat motif (allele 1) |
| gt_INS_GangSTR | GangSTR | Insert size mean (allele 1) |
| gt_STDERR_GangSTR | GangSTR | Insert size stddev (allele 1) |
| info_STUTTERUP_GangSTR | GangSTR | Stutter model - up prob |
| info_STUTTERDOWN_GangSTR | GangSTR | Stutter model - down prob |
| info_STUTTERP_GangSTR | GangSTR | Stutter model - p |
| gt_DP_GangSTR | GangSTR | Read Depth |
| gt_Q_GangSTR | GangSTR | Quality Score (posterior probability) |
| gt_ML_GangSTR | GangSTR | Maximum likelihood |
| gt_QEXP_1_GangSTR | GangSTR | Prob. of no expansion, 1 expanded allele, both expanded alleles (1) |
| gt_QEXP_2_GangSTR | GangSTR | Prob. of no expansion, 1 expanded allele, both expanded alleles (2) |
| gt_QEXP_3_GangSTR | GangSTR | Prob. of no expansion, 1 expanded allele, both expanded alleles (3) |
| gt_RC_1_GangSTR | GangSTR | Number of reads in enclosing class |
| gt_RC_2_GangSTR | GangSTR | Number of reads in spanning class |
| gt_RC_3_GangSTR | GangSTR | Number of reads in FRR class |
| gt_RC_4_GangSTR | GangSTR | Number of reads in bounding class |
| otherA_GangSTR | GangSTR | poly-T length predicted by GangSTR (allele 2) |
| kmer_3 | Jellyfish | Number of k-mer size = 3 |
| kmer_4 | Jellyfish | Number of k-mer size = 4 |
| kmer_5 | Jellyfish | Number of k-mer size = 5 |
| kmer_6 | Jellyfish | Number of k-mer size = 6 |
| kmer_7 | Jellyfish | Number of k-mer size = 7 |
| kmer_8 | Jellyfish | Number of k-mer size = 8 |
| kmer_9 | Jellyfish | Number of k-mer size = 9 |
| kmer_10 | Jellyfish | Number of k-mer size = 10 |
| kmer_11 | Jellyfish | Number of k-mer size = 11 |
| kmer_12 | Jellyfish | Number of k-mer size = 12 |
| kmer_13 | Jellyfish | Number of k-mer size = 13 |
| kmer_14 | Jellyfish | Number of k-mer size = 14 |
| kmer_15 | Jellyfish | Number of k-mer size = 15 |
| kmer_16 | Jellyfish | Number of k-mer size = 16 |
| kmer_17 | Jellyfish | Number of k-mer size = 17 |
| kmer_18 | Jellyfish | Number of k-mer size = 18 |
| kmer_19 | Jellyfish | Number of k-mer size = 19 |
| kmer_20 | Jellyfish | Number of k-mer size = 20 |
| kmer_21 | Jellyfish | Number of k-mer size = 21 |
| kmer_22 | Jellyfish | Number of k-mer size = 22 |
| kmer_23 | Jellyfish | Number of k-mer size = 23 |
| kmer_24 | Jellyfish | Number of k-mer size = 24 |

| | | |
|---|---|---|
| kmer_25 | Jellyfish | Number of k-mer size = 25 |
| kmer_26 | Jellyfish | Number of k-mer size = 26 |
| kmer_27 | Jellyfish | Number of k-mer size = 27 |
| kmer_28 | Jellyfish | Number of k-mer size = 28 |
| kmer_29 | Jellyfish | Number of k-mer size = 29 |
| kmer_30 | Jellyfish | Number of k-mer size = 30 |
| kmer_31 | Jellyfish | Number of k-mer size = 31 |
| kmer_32 | Jellyfish | Number of k-mer size = 32 |
| kmer_33 | Jellyfish | Number of k-mer size = 33 |
| kmer_34 | Jellyfish | Number of k-mer size = 34 |
| kmer_35 | Jellyfish | Number of k-mer size = 35 |
| kmer_36 | Jellyfish | Number of k-mer size = 36 |
| kmer_37 | Jellyfish | Number of k-mer size = 37 |
| kmer_38 | Jellyfish | Number of k-mer size = 38 |
| kmer_39 | Jellyfish | Number of k-mer size = 39 |
| kmer_40 | Jellyfish | Number of k-mer size = 40 |
| kmer_41 | Jellyfish | Number of k-mer size = 41 |
| kmer_42 | Jellyfish | Number of k-mer size = 42 |
| kmer_43 | Jellyfish | Number of k-mer size = 43 |
| kmer_44 | Jellyfish | Number of k-mer size = 44 |
| kmer_45 | Jellyfish | Number of k-mer size = 45 |
| kmer_46 | Jellyfish | Number of k-mer size = 46 |
| kmer_47 | Jellyfish | Number of k-mer size = 47 |
| kmer_48 | Jellyfish | Number of k-mer size = 48 |
| kmer_49 | Jellyfish | Number of k-mer size = 49 |
| kmer_50 | Jellyfish | Number of k-mer size = 50 |
| .pred_class | MLP | 6 TOMM40'523 genotype categories predicted using MLP |
| .pred_1 | MLP | Probabilities of class 1 (S/S) as predicted by the MLP |
| .pred_2 | MLP | Probabilities of class 2 (S/L) as predicted by the MLP |
| .pred_3 | MLP | Probabilities of class 3 (S/VL) as predicted by the MLP |
| .pred_4 | MLP | Probabilities of class 4 (L/L) as predicted by the MLP |
| .pred_5 | MLP | Probabilities of class 5 (L/VL) as predicted by the MLP |
| .pred_6 | MLP | Probabilities of class 6 (VL/VL) as predicted by the MLP |
| pred_S | MLP | Probabilities of another 3-class MLP model to predict class 1 (S-allele) |
| pred_L | MLP | Probabilities of another 3-class MLP model to predict class 2 (L-allele) |
| pred_VL | MLP | Probabilities of another 3-class MLP model to predict class 3 (VL-allele) |

**Supp. table S2. Description of ADRD traits tested**

| variable | family | description |
|---|---|---|
| cogn_global_lv | gaussian | Global cognitive function |
| cogng_demog_slope | gaussian | Cognitive decline |
| cogng_path_slope | gaussian | Cognitive resilience |
| amyloid_sqrt | gaussian | Beta-Amyloid |
| tangles_sqrt | gaussian | Tangle density |
| tdp_43_binary | binomial | TDP-43 |
| dlbany | binomial | Lewy Body disease |
| caa_4gp | ordinal | Cerebral amyloid angiopathy |
| cvda_4gp2 | ordinal | Cerebral Atherosclerosis |
| arteriol_scler | ordinal | Arteriolosclerosis |
| cogn_ep_lv | gaussian | Episodic memory |
| cogn_po_lv | gaussian | Visuospatial ability/perceptual orientation |
| cogn_ps_lv | gaussian | Perceptual speed |
| cogn_wo_lv | gaussian | Working memory |
| cogn_se_lv | gaussian | Semantic memory |

**Supp. table S3. Concordance of TOMM40-'523 genotypes and *APOE* ε4 (NLW-test samples only)**

| *APOE* | 523-S/S | 523-S/L or L/L or L/VL | 523-S/VL or VL/VL |
|---|---|---|---|
| *PCR-derived '523* | | | |
| ε4- | 101 | 3 | 299 |
| ε4+ | 1 | 137 | 9 |
| *WGS-derived '523* | | | |
| ε4- | 95 | 20 | 288 |
| ε4+ | 0 | 138 | 9 |

**Supp. table S4. Concordance of TOMM40-'523 genotypes and *APOE* ε4 (NLB-test samples only)**

| *APOE* | 523-S/S | 523-S/L or L/L or L/VL | 523-S/VL or VL/VL |
|---|---|---|---|
| *PCR-derived '523* | | | |
| ε4- | 7 | 2 | 9 |
| ε4+ | 4 | 6 | 4 |
| *WGS-derived '523* | | | |
| ε4- | 6 | 3 | 9 |
| ε4+ | 4 | 7 | 3 |
